# Simply saliva: stability of SARS-CoV-2 detection negates the need for expensive collection devices

**DOI:** 10.1101/2020.08.03.20165233

**Authors:** Isabel M. Ott, Madison S. Strine, Anne E. Watkins, Maikel Boot, Chaney C. Kalinich, Christina A. Harden, Chantal B.F. Vogels, Arnau Casanovas-Massana, Adam J. Moore, M. Catherine Muenker, Maura Nakahata, Maria Tokuyama, Allison Nelson, John Fournier, Santos Bermejo, Melissa Campbell, Rupak Datta, the Yale IMPACT Research team, Charles S. Dela Cruz, Shelli F. Farhadian, Albert I. Ko, Akiko Iwasaki, Nathan D. Grubaugh, Craig B. Wilen, Anne L. Wyllie

## Abstract

Most currently approved strategies for the collection of saliva for COVID-19 diagnostics require specialized tubes containing buffers promoted for the stabilization of SARS-CoV-2 RNA and virus inactivation. Yet many of these are expensive, in limited supply, and not necessarily validated specifically for viral RNA. While saliva is a promising sample type as it can be reliably self-collected for the sensitive detection of SARS-CoV-2, the expense and availability of these collection tubes are prohibitive to mass testing efforts. Therefore, we investigated the stability of SARS-CoV-2 RNA and infectious virus detection from saliva without supplementation. We tested RNA stability over extended periods of time (2-25 days) and at temperatures representing at-home storage and elevated temperatures which might be experienced when cold chain transport may be unavailable. We found SARS-CoV-2 RNA in saliva from infected individuals is stable at 4°C, room temperature (∼19°C), and 30°C for prolonged periods and found limited evidence for viral replication in stored saliva samples. This work demonstrates that expensive saliva collection options involving RNA stabilization and virus inactivation buffers are not always needed, permitting the use of cheaper collection options. Affordable testing methods are urgently needed to meet current testing demands and for continued surveillance in reopening strategies.

## Background

Despite an increase in diagnostic testing capacity for SARS-CoV-2, in many countries, including the United States, testing is still inadequate to slow the COVID-19 pandemic. Many people still do not have access to SARS-CoV-2 tests, and some that do still experience long delays in receiving results due to imbalance between supply and demand at large testing centers. The demand for testing will only increase with the reopening of many schools, colleges, and workplaces. Ideally, specialized population surveillance-oriented testing would (***1***) require minimal diversion of resources from clinical diagnostic testing, (***2***) be affordable and scalable, and (***3***) allow for rapid and reliable identification of virus presence for asymptomatic or subclinical infections. Thus, simplifying the sample collection and testing workflow is critical.

Collecting saliva for SARS-CoV-2 detection is one of the simple solutions needed to massively expand testing. We and others have shown that saliva is a sensitive source for SARS-CoV-2 detection^1–3^. Perhaps of equal importance, saliva collection is non-invasive, can be reliably done at home without trained health professions, and does not rely on a sometimes limited supply of swabs. However, the only saliva-based mass testing strategies currently approved by the U.S. Food and Drug Administration (FDA) require specialized collection tubes containing stabilization and/or inactivation buffers that are costly with unreliable availability. Moreover, as saliva continues to gain popularity as a potential specimen to aid testing demands, unlike traditional swab-based methods, standardized collection methods have not been defined. Additionally, when true saliva is not collected (*e*.*g*. contains sputum), which can happen with COVID-19 inpatients when saliva is difficult to produce, the sample type can be difficult to pipette. Combined with untested concerns regarding SARS-CoV-2 RNA stability in saliva, supplements to reduce degradation and improve sample processing have become common. Prior work from saliva samples, however, has indicated that some buffers optimized for host nucleic acid stabilization may actually inhibit viral RNA detection,^4–6^ particularly in extraction-free PCR workflows.^7^ Thus, if true saliva - which is relatively easy to pipette - is being tested, the utility of collecting saliva in expensive tubes containing purported stabilization buffers comes into question.

To explore the viability of broadly deploying affordable saliva-based surveillance approaches^8^, we characterized SARS-CoV-2 RNA stability and virus infectivity from saliva samples stored in widely available, sterile, nuclease-free laboratory plastic (polypropylene) tubes. We found stable detection of SARS-CoV-2 RNA in saliva samples at a range of temperatures and for prolonged periods, supporting the potential for inexpensive and simple saliva collection.

## Results

Saliva collected from COVID-19 inpatients and healthcare workers using sterile collection tubes^2^ was used to evaluate the temporal stability of SARS-CoV-2 RNA at different holding temperatures (-80°C, 4°C, ∼19°C, 30°C) without using nucleic acid preservatives. Importantly, we found that SARS-CoV-2 RNA from saliva was consistently detected at similar levels regardless of the holding time and temperatures tested. Following RNA extraction^9^ and RT-qPCR^10^ testing for SARS-CoV-2 on the day of saliva collection^2^, the remaining sample volumes (n=20) were aliquoted and stored at -80°C, room temperature (recorded as ∼19°C) and 30°C. Whether tested on day of collection or after storage at - 80°C freeze/thaw, room temperature (5 days), and 30°C (3 days), RT-qPCR cycle threshold (Ct) values for N1 were not significantly different (**Figure 1A**). Following the freeze/thaw cycle or storage at room temperature, we observed Ct decreases of 1.058 (95% CI: -2.289, 0.141) and 0.960 (95% CI: -2.219,0.266), respectively; however the strength of this effect was low. A similar effect was seen following incubation at 30°C with an increase of Ct 0.973 (95% CI: −0.252, 2.197). Moreover, SARS-CoV-2 RNA remained relatively stable in saliva samples left for up to 25 days at room temperature (∼19°C; Ct increase of 0.027, 95% CI: −0.019, 0.071) (**Figure 1B**). This finding is in line with a recent study also reporting on the stability of SARS-CoV-2 RNA in saliva at room temperature for up to 7 days^6^. Regardless of the starting Ct value (and therefore viral load), this prolonged stability of SARS-CoV-2 RNA was also observed when samples were stored for longer periods at -80°C (max. 92 days), 4°C (max. 21 days), and 30°C (max. 16 days) (**Supplemental Figure 1**).

**Figure 1.**
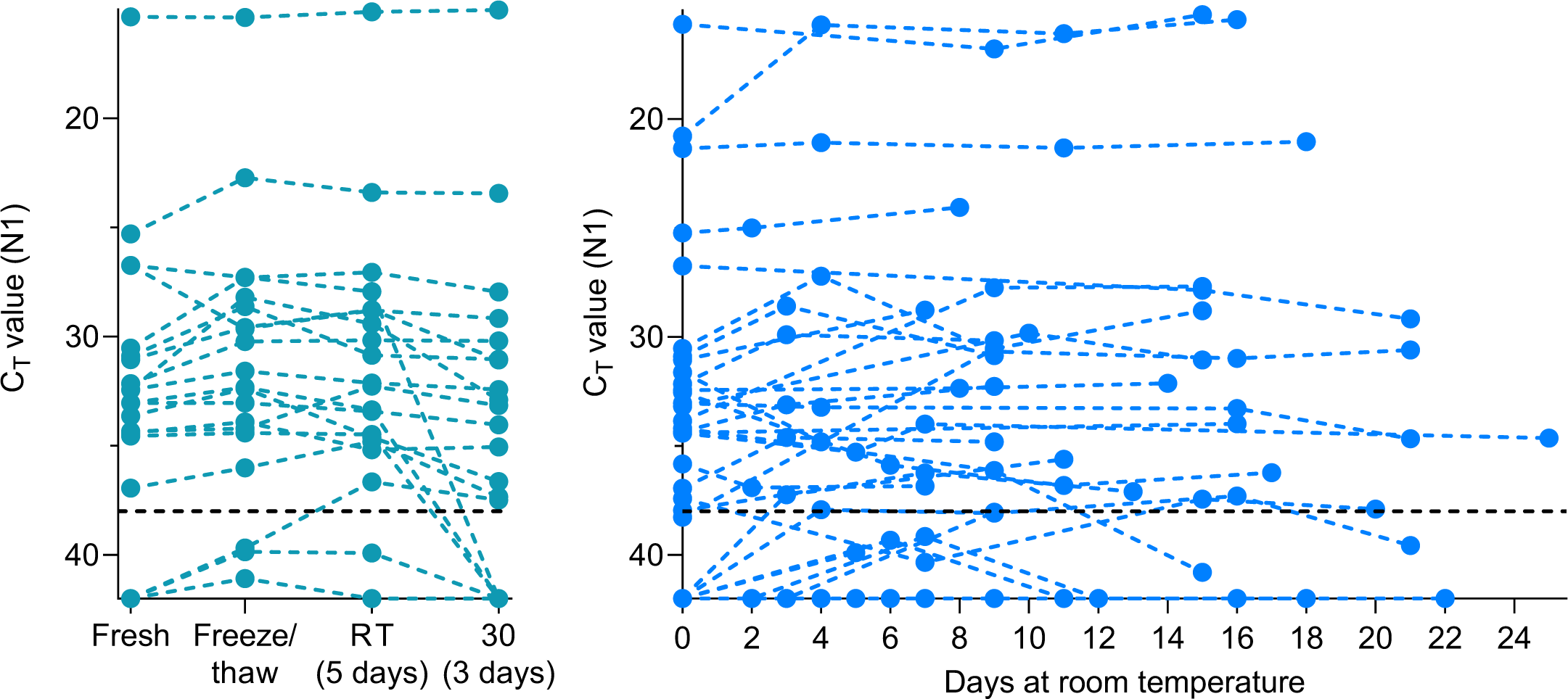
Stability of SARS-CoV-2 RNA detection in saliva. SARS-CoV-2 RNA detection in (A) saliva (n=20) on day of sample collection (fresh) or after storage at -80°C, 30°C for 3 days or room temperature (RT, recorded as ∼19°C) for 5 days. The detection of RNA remained stable regardless of starting Ct value (Pearson’s r = −0.085, *p* = 0.518). At room temperature (B), detection remained stable for up to 25 days. Ct values from the same sample in different conditions are connected by a dotted line. The black dashed line represents Ct 38 which we applied as the cut-off to determine sample positivity. Samples that remained not detected (ND) after 45 cycles are depicted as Ct 42.

Interestingly, while SARS-CoV-2 RNA from saliva remained stable over time, we observed a decrease in human *RNAse P* (RP) at higher temperatures (room temperature, Ct +1.837, 95% CI: 0.468, 3.188; 30°C, Ct +3.526, 95% CI: 1.750, 5.349; **Supplemental Figure 2**), with the change in concentration greater than that observed for SARS-CoV-2 RNA (**Supplemental Figure 3**). Thus, our data indicates that while human RNA from saliva degrades without stabilization buffers, SARS-CoV-2 RNA remains protected even at warm temperatures suited for nuclease activity.

As saliva has been shown to have antiviral properties^11,12^, we explored the infectiousness of SARS-CoV-2 present in saliva samples. We inoculated Vero-E6 cells with saliva samples of higher virus RNA titers (**Supplemental Figure 4**), as others have shown that SARS-CoV-2 isolation is uncommon at low virus RNA titers^13–16^. By 72 hours post-inoculation, five of the 43 (11.6%) saliva samples cultured exhibited a reduction in Ct values when tested by RT-qPCR (-4.41, -4.3, -3.86, -3.49 and -2.66, **Figure 2**). While these findings suggest an increase in the number of SARS-CoV-2 RNA copies by 72 hours, this may not definitively demonstrate active viral replication. For instance, Ct reductions could also likely result from sampling artifacts or assay variations (disparities in inoculation, RNA extraction, and RT-qPCR). To determine whether this amplification resulted from active viral replication, we performed plaque assays with cellular lysate from 72 hours post-inoculation. Interestingly, no plaque forming units (PFU) could be visualized after 48 hours post-infection. This may suggest that the increase in SARS-CoV-2 genome copies identified by RT-qPCR may not have resulted from viral replication, that infectious virus falls below the limit of detection (100 PFU/mL), or possibly that components of saliva inhibit active viral particle production and release. A similar result was observed when attempting to plaque virus from the colon^17^, despite studies showing that SARS-CoV-2 infected gut enterocytes^18^.

**Figure 2.**
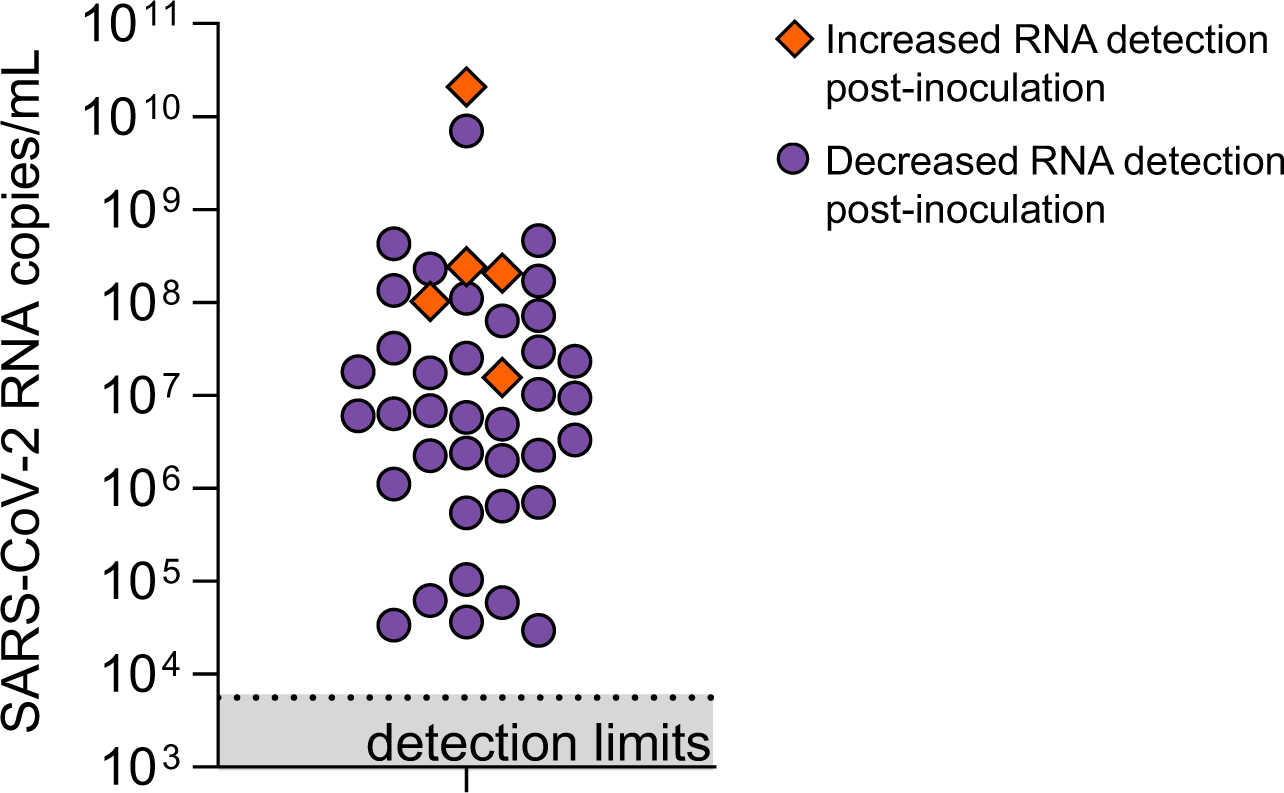
Viral load of saliva samples tested for infectious SARS-CoV-2. Starting viral load (calculated from RT-qPCR detection of N1) of saliva samples incubated with Vero-E6 cells for 72 hours. Orange diamonds depict samples in which a reduction in Ct value of >2 at 72 hours post-inoculation was observed as compared to 1 hour post-inoculation. Plaque assays with the cellular lysate from 72 hours post-inoculation however, resulted in no plaque forming units (PFU) after 48 hours post-infection.

## Discussion

Inexpensive saliva-based SARS-CoV-2 testing methods are urgently needed to help reach the capacity needed to safely reopen schools and workplaces. We demonstrate the stability of SARS-CoV-2 RNA detection for prolonged periods in a variety of settings, which indicates that saliva can be simply collected without the need of expensive additives. With commercial tubes being promoted as specialized for the collection of saliva and stabilization of SARS-CoV-2 RNA costing over $7 per tube (**Table 1**), these costs can be prohibitive to mass testing efforts. Moreover, previous studies have demonstrated the ease in which saliva can be collected in simple, wide-mouth containers^2,9,19^ and that buffers marketed for RNA stabilization may be detrimental to SARS-CoV-2 detection^6^. Without the need for RNA stabilization and with limited evidence of viral replication in saliva samples, simple, sterile, nuclease-free plastic containers are affordable alternatives to making testing accessible throughout the country. SARS-CoV-2 stability at both room temperature and 30°C permits more affordable collection and transport strategies without a need for expensive cooling strategies. Without the requirement for cold chain handling, this also facilitates the implementation of saliva testing in regions or countries with limited resources. Thus, collection of saliva in simple, sterile, nuclease-free tubes, negating the high costs associated with specialized collection devices, is one of the keys to meet mass testing demands.

**Table 1.**
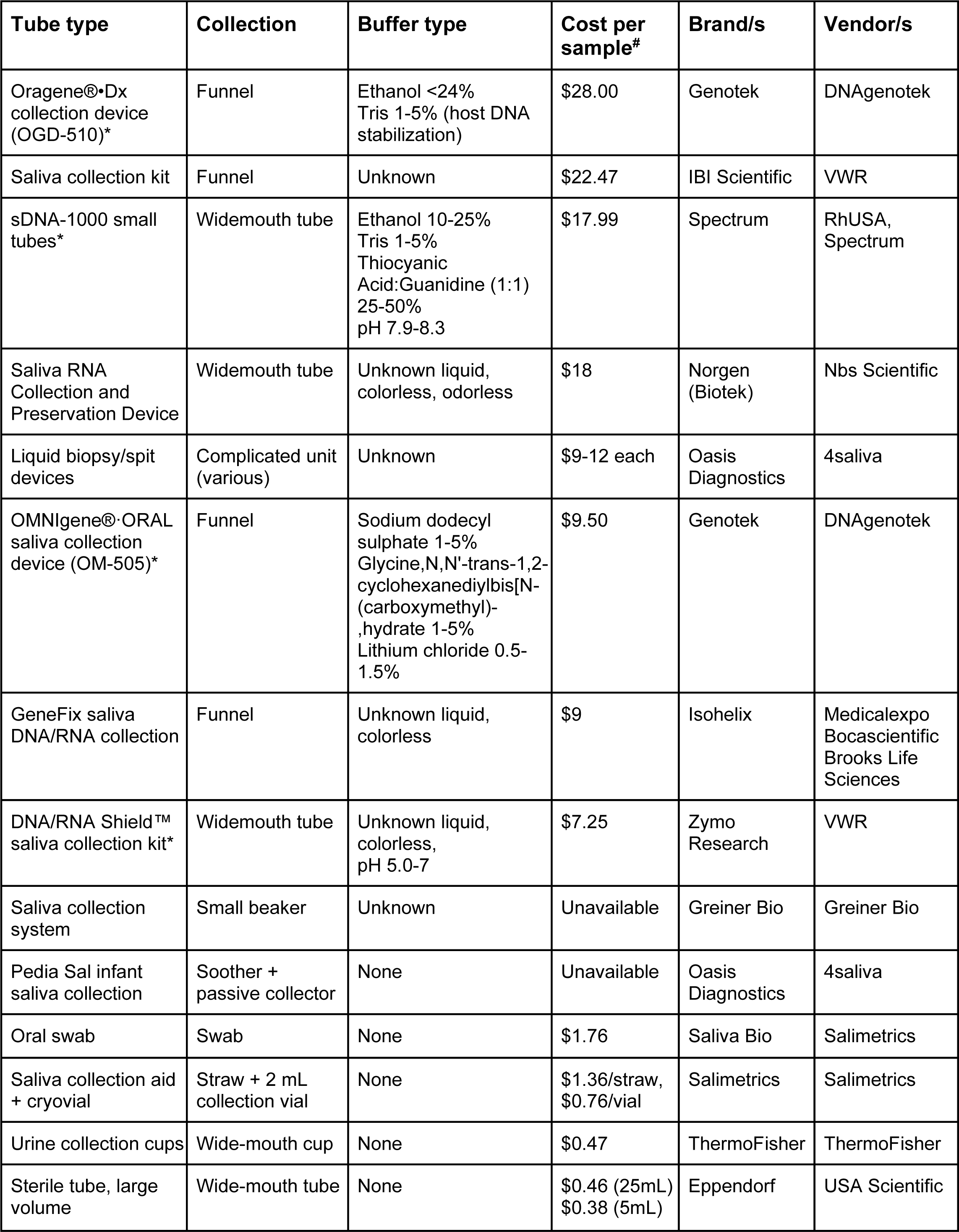
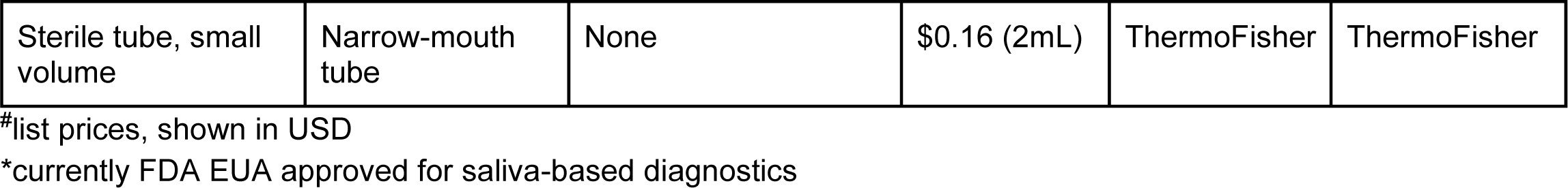
Possible saliva collection devices: type, approach and list price (USD) per sample.

## Methods

### RNA extraction and SARS-CoV-2 detection

RNA was extracted from saliva samples^9^ collected from COVID-19 inpatients and healthcare workers at the Yale-New Haven Hospital (Yale Human Research Protection Program Institutional Review Boards FWA00002571, Protocol ID. 2000027690)^2^. RNA templates were tested by RT-qPCR for SARS-CoV-2 RNA (N1)^10^ on day of collection (∼12 hours post collection) and at various time points after the storage of the remaining, unsupplemented samples at temperatures of -80°C, -20°C, +4°C, 19°C, or 30°C.

### Cell culture

Vero-E6 cells (ATCC) were cultured in Dulbecco’s Modified Eagle Medium (Gibco) supplemented with 10% heat-inactivated fetal bovine serum (VWR), 1% Penicillin/Streptomycin (Gibco), 100 µg/mL gentamicin (Gibco), and 0.5 µg/mL amphotericin B (Fisher Scientific). All cells were incubated at 37°C and 5% CO2. All cell culture experiments were performed in a biosafety level 3 laboratory at Yale University and approved by the Yale University Biosafety Committee.

### Saliva inoculation and serial passaging in Vero-E6 cells

Saliva samples were diluted 1:1 in 1X Dulbecco’s PBS (Gibco). Diluted saliva samples were incubated for one hour at 37°C with 2.5×10^5^ Vero-E6 cells in a 24-well plate (Corning). Unbound virus was aspirated and the media were replaced. Infected Vero-E6 cells were frozen at -80°C at 1 and 72 hours post-inoculation. Prior to RNA extraction^9^ and RT-qPCR detection of SARS-CoV-2 RNA^10^ the Vero-E6 cells from 1 and 72 hours post-inoculation were thawed at room temperature and further lysed by diluting 1:3 in MagMax Binding Solution (ThermoFisher). RNA was extracted from the two timepoints and tested in RT-qPCR for SARS-CoV-2 N1. We interpreted a Ct reduction >2 as a difference which could potentially be explained by viral replication during the two timepoints.

### Plaque assay

Vero-E6 cells were seeded at 4×10^5^ cells/well in 12-well plates. The following day, media were removed and replaced with 100ul of 10-fold serial dilutions of thawed 72 hour post-inoculation saliva samples. Plates were incubated at 37°C for 1 hour with gentle rocking every 10 mins. Unbound inocula was aspirated from each well and overlay media (DMEM, 2% FBS, 0.6% Avicel RC-581 (DuPont)) was added to each well. At 48 hours post-infection, plates were fixed with 5-10% formaldehyde for 30 min then stained with crystal violet solution (0.5% crystal violet in 20% ethanol) for 30 mins. Crystal violet solution was then aspirated, and plates were washed in tap water to visualize plaques.

### Statistical analyses

We fit a linear regression to the experimental stability data to model the change in Ct values of positive samples following stability conditions using the equation below. Let dct be the change in Ct value from fresh testing following each storage condition and let condition be the categorical storage condition (e.g. freeze/thaw, room temperature, 30°C, etc).

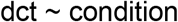

Robust confidence intervals were simulated from this model using the mvrnorm, in the R package “MASS”, and quantile functions. This regression was also used to model the effect of prolonged storage in stability conditions on RP.

For extended timepoint analyses of N1 we used a linear mixed effects model to predict the change in Ct values of positive samples under each stability condition for greater durations of time using the equation below. Let timepoint be the number of days under stability conditions and let sample be the patient number.

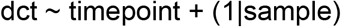

Confidence intervals were computed for this model using confint.merMod, in the R package “lme4”.

Further statistical analyses were conducted in GraphPad Prism 8.0.0 as described in the text and figure legends.

### Yale IMPACT Research Team authors (in alphabetical order)

Kelly Anastasio, Michael H. Askenase, Maria Batsu, Sean Bickerton, Kristina Brower, Molly L. Bucklin, Staci Cahill, Yiyun Cao, Edward Courchaine, Giuseppe DeIuliis, Rebecca Earnest, Renata Filler, Bertie Geng, Benjamin Goldman-Israelow, Ryan Handoko, William Khoury-Hanold, Daniel Kim, Lynda Knaggs, Maxine Kuang, Eriko Kudo, Sarah Lapidus, Joseph Lim, Melissa Linehan, Peiwen Lu, Alice Lu-Culligan, Anjelica Martin, Irene Matos, David McDonald, Maksym Minasyan, Nida Naushad, Jessica Nouws, Abeer Obaid, Camila Odio, Ji Eun Oh, Saad Omer, Annsea Park, Hong-Jai Park, Xiaohua Peng, Mary Petrone, Sarah Prophet, Tyler Rice, Kadi-Ann Rose, Lorenzo Sewanan, Lokesh Sharma, Denise Shepard, Mikhail Smolgovsky, Nicole Sonnert, Yvette Strong, Codruta Todeasa, Jordan Valdez, Sofia Velazquez, Arvind Venkataraman, Pavithra Vijayakumar, Elizabeth B. White, Yexin Yang.

## Data Availability

The data that support the findings of this study are available from the corresponding author, [ALW], upon reasonable request.

## Acknowledgements

We gratefully acknowledge the study participants for their time and commitment to the study. We thank all members of the clinical team at Yale-New Haven Hospital for their dedication and work which made this study possible. We also thank S. Taylor and P. Jack for technical discussions.

## Funding

This study was funded by the Huffman Family Donor Advised Fund, Fast Grant funding support from the Emergent Ventures at the Mercatus Center, George Mason University, the Yale Institute for Global Health, and the Beatrice Kleinberg Neuwirth Fund. CBFV is supported by NWO Rubicon 019.181EN.004. CBW is supported by NIH K08 AI128043, the Burroughs Wellcome Fund, and the Ludwig Family Foundation.

## Competing interests

ALW has received research funding through grants from Pfizer to Yale and has received consulting fees for participation in advisory boards for Pfizer.

## Supplemental Figures

**Supplemental Figure 1.**
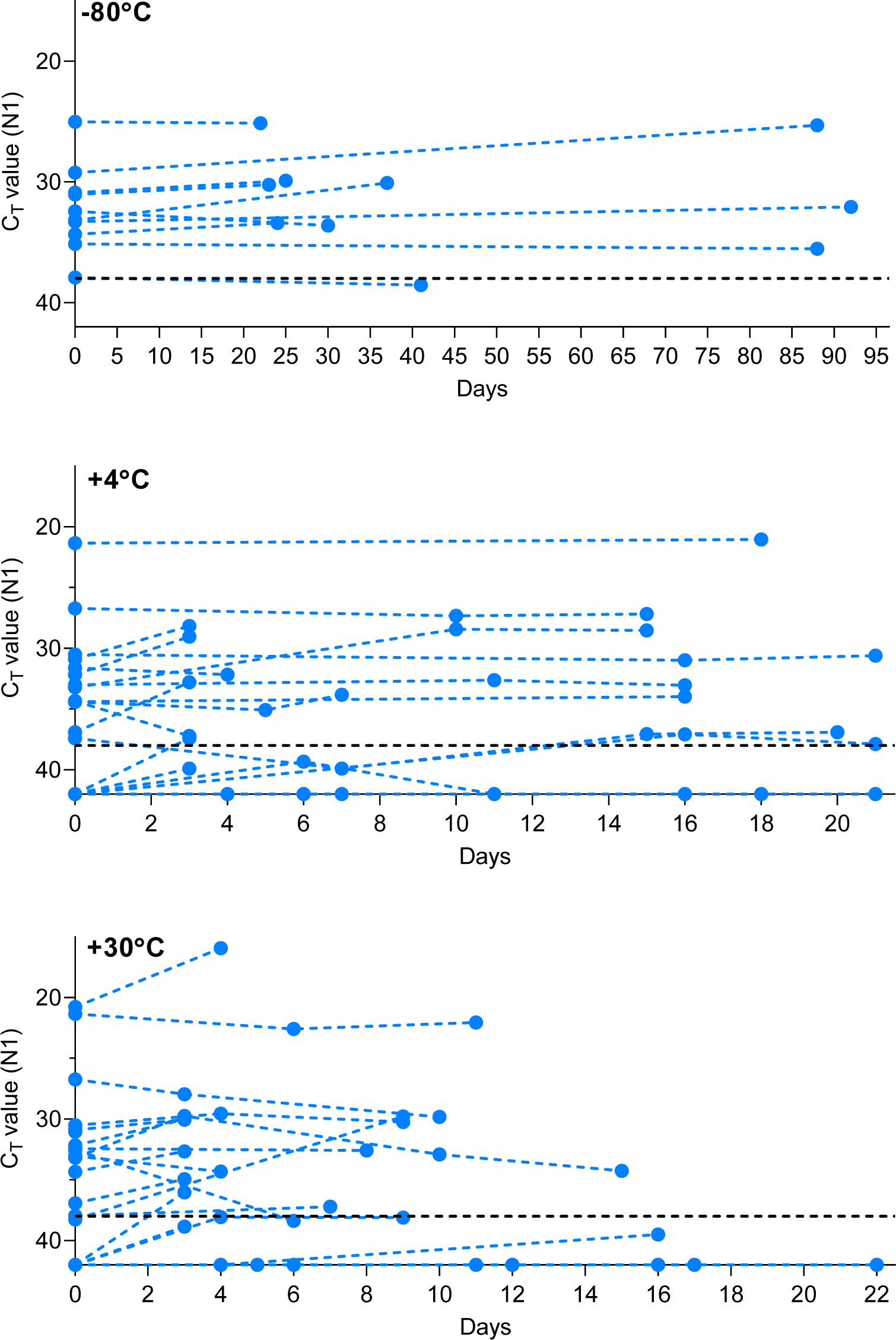
Stability of SARS-CoV-2 RNA detection in saliva. SARS-CoV-2 RNA detection in saliva on day of sample collection (0) or after prolonged storage at -80°C, 4°C or 30°C. Ct values from the same original sample are connected by a dotted line. The -80°C and 4°C conditions were found to have a weakly beneficial effect on signal detection by the mixed effects model, while the 30°C condition resulted in a slight increase in Ct. The -80°C storage alone did not cross zero suggesting a mildly stronger effect than the other conditions (95% CI: −0.038, −0.010). The black dashed line represents Ct 38 which we applied as the cut-off to determine sample positivity. Samples that remained not detected (ND) after 45 cycles are depicted as Ct 42.

**Supplemental Figure 2.**
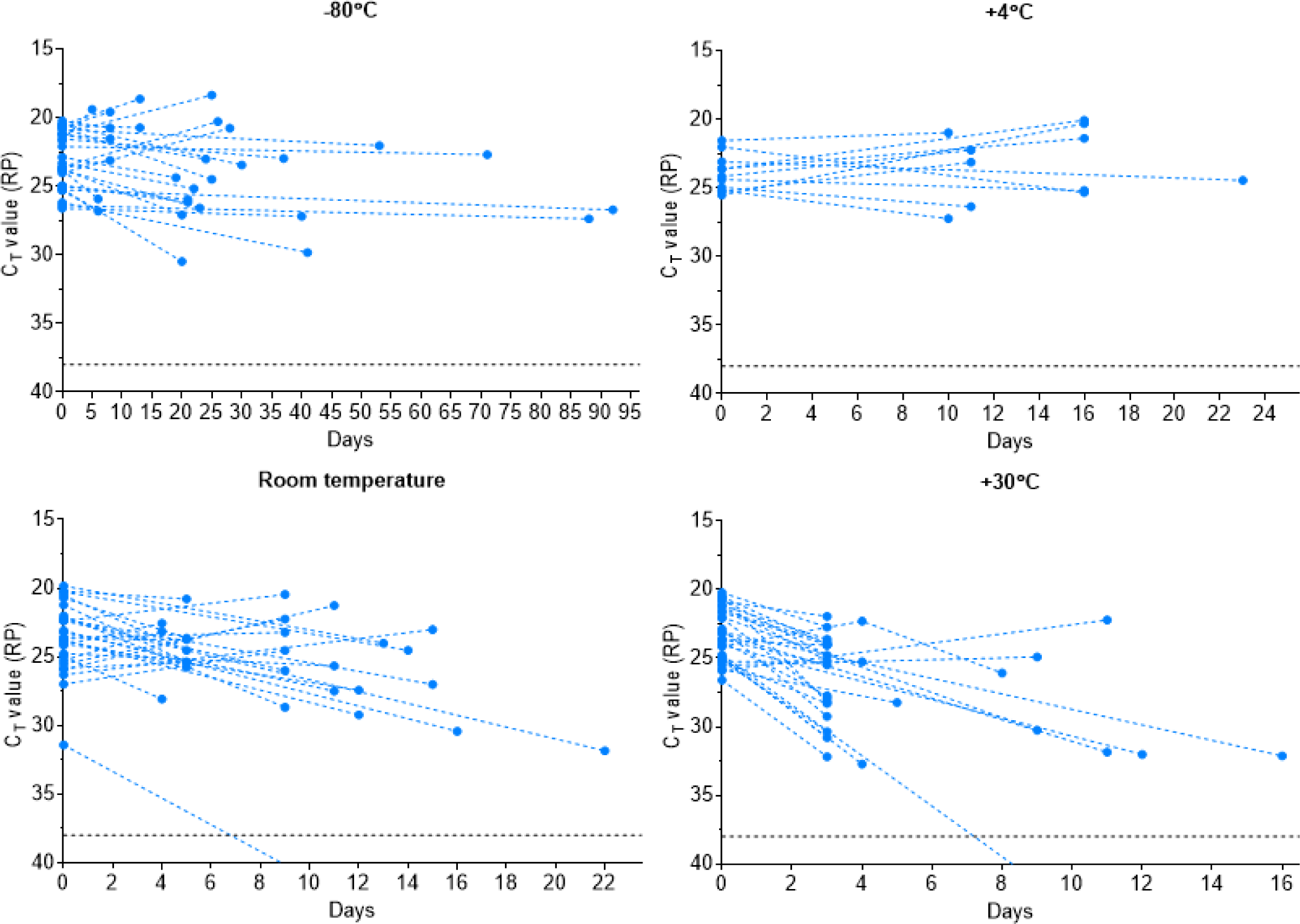
Detection of human *RNAse P* (RP) declines over time when stored in saliva in warmer conditions. Detection of human RP in saliva on day of collection (0) or after prolonged storage at -80°C, 4°C, room temperature (∼19°C) or 30°C. Ct values from the same original sample are connected by a dotted line. Prolonged storage at -80°C and 4°C had minimal effect on RP detection with Ct changes of 0.832 (95% CI: - 0.402, 2.038) and −0.315 (95% CI: -2.336, 1.687), respectively. However, storage at room temperature (Ct +1.837, 95% CI: 0.468, 3.188) and 30°C (Ct +3.526, 95% CI: 1.750, 5.349) was detrimental to RP, exhibiting a more substantial decrease in signal at these warmer conditions. The black dashed line represents Ct 38 which we applied as the cut-off to determine sample positivity. Samples that remained not detected (ND) after 45 cycles are below the y-axis limit.

**Supplemental Figure 3.**
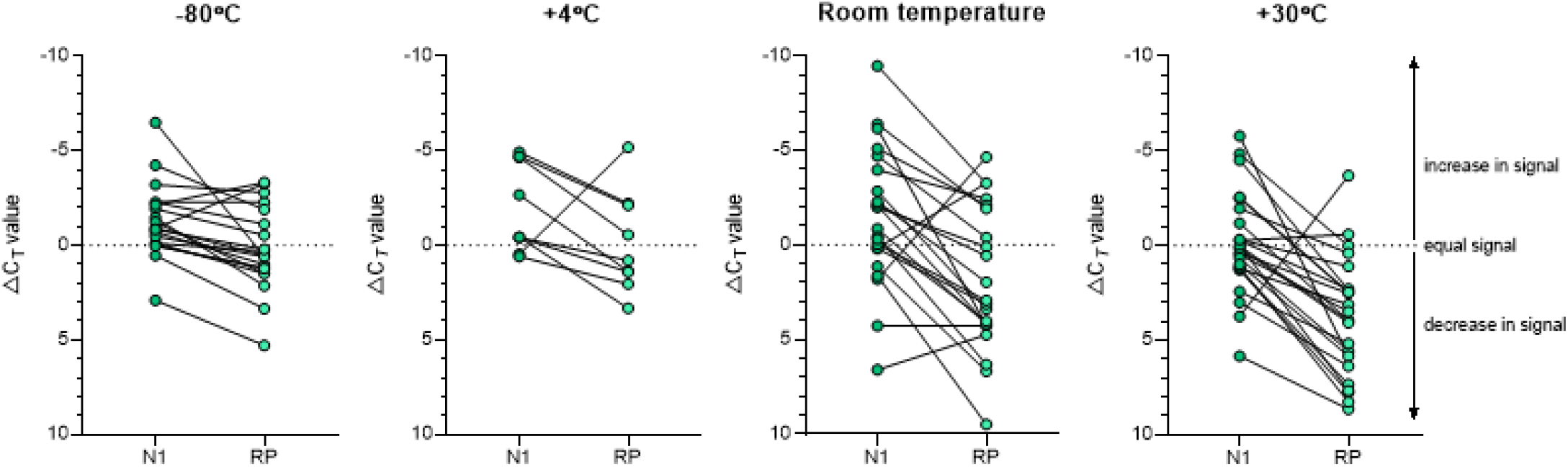
Detection of SARS-CoV-2 RNA (N1) in saliva remained more stable over time than human *RNAse P* (RP). Delta Ct was calculated as the difference in Ct value from the day of saliva collection and after storage at -80°C, 4°C, room temperature (∼19°C) or 30°C. Delta Ct values from the same sample are joined by a solid line. While the change in detection of SARS-CoV-2 N1 and RP was similar in saliva samples stored at 4°C (Wilcoxon signed rank test, *p* = 0.129), a greater difference was observed between the change in N1 and RP for samples stored at -80°C (*p* = 0.001), room temperature (*p* = 0.001) and 30°C (*p* < 0.0001).

**Supplemental Figure 4.**
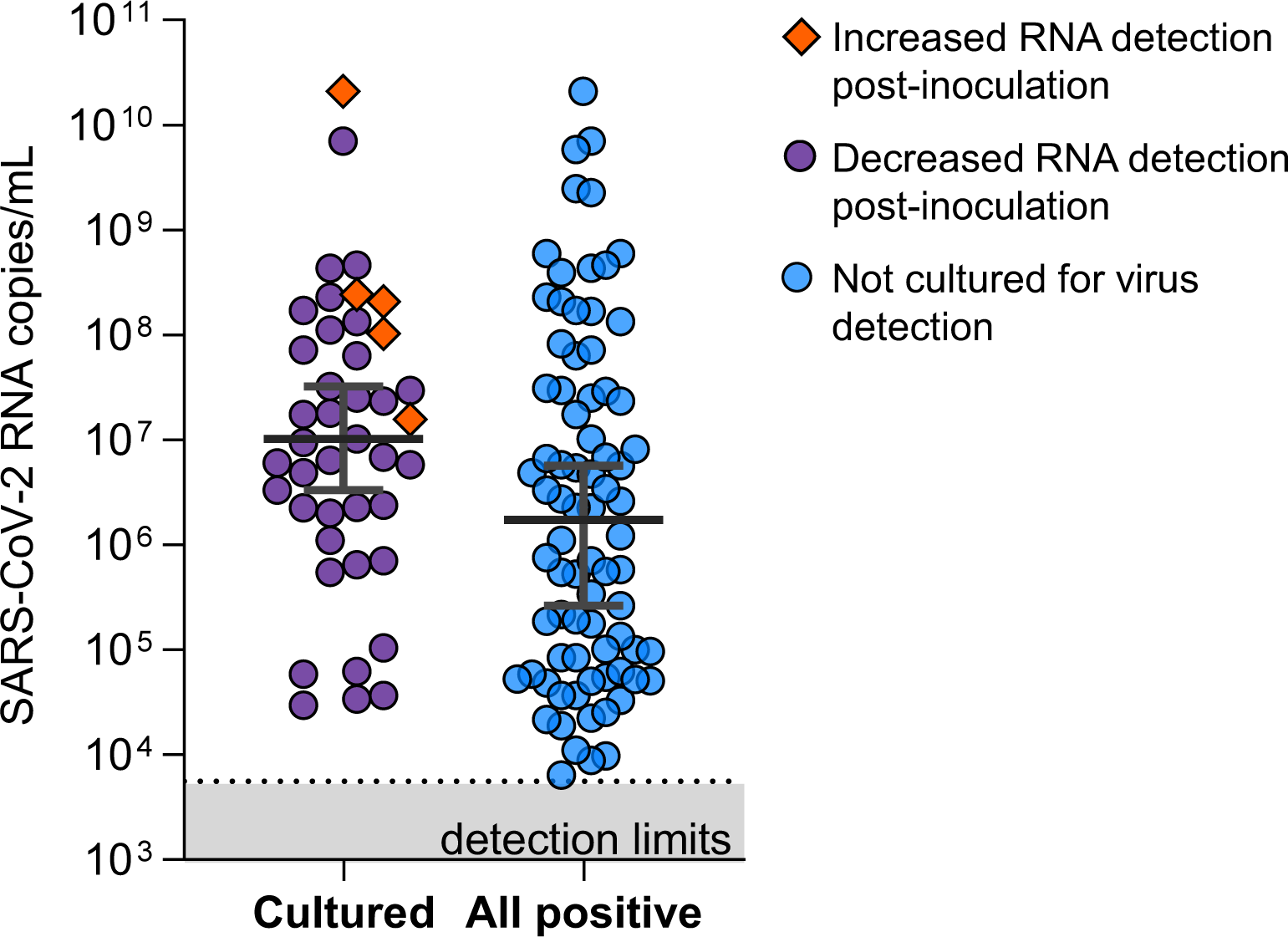
Saliva samples of relatively high viral load were cultured to evaluate the infectiousness of SARS-CoV-2 in saliva. Saliva samples cultured on Vero-E6 to test for infectious virus were of higher SARS-CoV-2 RNA (N1) load as compared to the overall saliva samples collected by Yale IMPACT^2^ which tested positive for SARS-CoV-2 (Mann-Whitney, *p* = 0.0136). Orange diamonds denote samples in which we observed an increase in viral RNA detection 72 hours post-inoculation.

## Notes

### Competing Interest Statement

The authors have declared no competing interest.

### Author Declarations

All study participants were enrolled and sampled in accordance to the Yale University HIC-approved protocol #2000027690. Demographics, clinical data and samples were collected after the study participant had acknowledged that they had understood the study protocol and signed the informed consent. All participant information and samples were collected in association with non-individually identifiable study identifiers.

